# Augmenting Large Language Models and Retrieval-Augmented Generation with an Evidence-Based Medicine-Enabled Agent System

**DOI:** 10.1101/2025.10.17.25338266

**Authors:** Yi Yu, Lingli Li, Yaqin Li

**Author notes:** **Correspondence:** (Yanqin Li), (Lingli Li).

## Abstract

**Importanc:** Large language models (LLMs) with retrieval-augmented generation (RAG) show promise for clinical decision support. However, their application is constrained by limited & outdated vector databases, suboptimal evidence retrieval and poor contextual continuity.

**Objective:** To develop and evaluate a novel LLM-based agent that integrates Evidence-Based Medicine (EBM) principles and contextual conversation capabilities in answering clinical questions.

**Design, Setting and Participants:** The agent for clinical decision making was developed and evaluated between July 1, 2024, and July 31, 2025. The system incorporated an EBM-enabled workflow, a memory module and Thought-Action-Observation (TAO) loops. Evaluation 1 assessed the system’s performance on 150 initial clinical questions across 15 cancer types. Evaluation 2 involved 45 multi-turn dialogue tasks (across 3 types). Baselines were state-of-the-art traditional RAG method and commercial LLMs with plugins. All generated responses across both evaluations were independently rated by 3 experts with over five years of clinical experience. The study was performed at West China Medical Center.

**Main Outcomes and Measures:** Each response in evaluation 1 was classified into one of three predefined categories—correct, inaccurate, or wrong. As for evaluation 2, tasks were deemed successful when previous conversation is remembered and answer is correct, otherwise the task was considered unsuccessful.

**Results:** In evaluation 1, EBMChat generated the highest proportion of accurate responses (89% vs 78% for the best baseline method). The superior performance of EBMChat was associated with its ability to retrieve optimal evidence, demonstrated by significantly higher evidence hierarchy (100% vs 17.5% RCT-level or above), stricter evidence timeliness (within 5 years vs from the 1980s onwards), and more comprehensive retrieval (median of 693 vs 267 items/question). Regarding evaluation 2, EBMChat successfully completed 93% of the tasks. In contrast, GPT-4.1 with plugins (Web Search) achieved a success rate of only 31%. This performance gap was attributed to EBM-enabled workflow, memory module and TAO loops, which ensure robust contextual conversation capabilities.

**Conclusion and Relevance:** EBMChat identifies appropriate evidence by effectively balancing timeliness, hierarchy, and relevance. Meanwhile, its enhanced conversational capabilities facilitate the preservation of contextual data, enabling users to explore clinical problems more deeply or comprehensively in multi-turn dialogues. Our findings underscore that the effective promotion of clinical practice by AI requires deeper integration of core medical principles into the technology itself, rather than direct application of general-purpose AI tools.

## Introduction

In recent years, considerable improvement in clinical decision-making has been validated through the combination of large language models (LLMs) and retrieval augmented generation (RAG)^1-5^. The RAG framework functions by vectorizing clinical queries and external medical documents, retrieving the most relevant evidence from a vector database, and leveraging LLMs to generate responses grounded in this evidence, thereby mitigating hallucinations^6-8^. Furthermore, LLM-based agents exhibit the potential to significantly expand these applications. Built upon LLMs, these agents can integrate external tools and perform goal-oriented reasoning and action to tackle clinical complex tasks^9-11^. For example, Ferber et al^12^ reported an LLM-based agent integrated with multimodal precision oncology tools (including RAG) to support personalized clinical chatbot in oncology, which substantially improved diagnostic accuracy.

Although RAG and agent technologies have facilitated the development of clinical decision-making, three challenges remain. Firstly, the limited scale of medical vector databases constrains the generalizability of RAG applications. Many existing efforts rely on self-collected databases, which often lack diversity in disease coverage, temporal span, and types of literature (e.g., only including guidelines)^13^. Although publicly available sources, such as datasets^14^ from MEDRAG, provide relatively comprehensive vector datasets, they lack real-time updates, resulting in the retrieval of outdated knowledge^15^. Secondly, RAG technology may not acquire the best evidence for answer generation. Actually, RAG selects the evidence by sorting vector database according to its relevance to the question. As for clinical decision-making, the relevance is an important but not the only evaluation metric for evidence screening. The neglect of critical factors such as evidence timeliness and hierarchy can lead to unreliable response generation^6^. Thirdly, multi-turn dialogue, instead of single-turn, is often essential for addressing clinical problems in real-world clinical practice^16-19^. However, traditional RAG frameworks typically exhibit limitations in retaining contextual information across multiple interactions, constraining their efficacy in multi-turn clinical interactions.

Evidence-based medicine (EBM) plays a crucial role in clinical decision-making through reasonable question conversion and evidence collection^20-22^. Inspired by the problem-solving ways of EBM, we have developed a new LLM-based agent, named EBMChat, to extend the application of LLMs and RAG. Through EBM-enabled workflow, EBMChat can step-by-step convert questions into queries, optimize queries, search databases, evaluate evidence, finally answer questions. In this way, EBMChat is capable of applying most suitable evidence from well-recognized databases^23,24^, simultaneously ensuring high timeliness, appropriate evidence hierarchy and the relevance with questions. Simultaneously, a memory module is designed to retain contextual information generated throughout the EBM workflow, including medical Boolean queries, search metadata, and details of the selected evidence. In subsequent clinical dialogues, this module and agent’s reasoning ability work in synergy to ensure user interactivity and problem-solving efficiency. We evaluated EBMChat using 150 clinical questions spanning 15 common cancer types. Based on rigorous manual evaluation by three clinical experts, the accuracy of EBMChat outputs surpasses state-of-the-art traditional RAG, as well as web search methods (89% vs 78%). In multi-turn dialogue tasks, EBMChat also significantly outperformed a leading commercial LLM (93% vs 31% task success rate).

## Method

### Design of EBMChat

The EBMChat agent is composed of five fundamental components: the LLM, toolset, prompts, executor and memory module. LLMs serves as the planner and brain of the agent to make decisions and select the appropriate tools for the action. We have utilized OpenAI’s GPT-4.1^25^ as the LLM, employing its advanced abilities in reasoning and planning to handle complicated tasks beyond text generation. Additionally, prompts can guide and standardize the behavior of LLMs^26^. Regarding the toolset, it consists of five sections: Query, Search, Evidence, Answer, and Chat (**Figure 1a**). Detailed descriptions of representative tools and prompts are provided in the **eMethods**.

**Figure 1.**
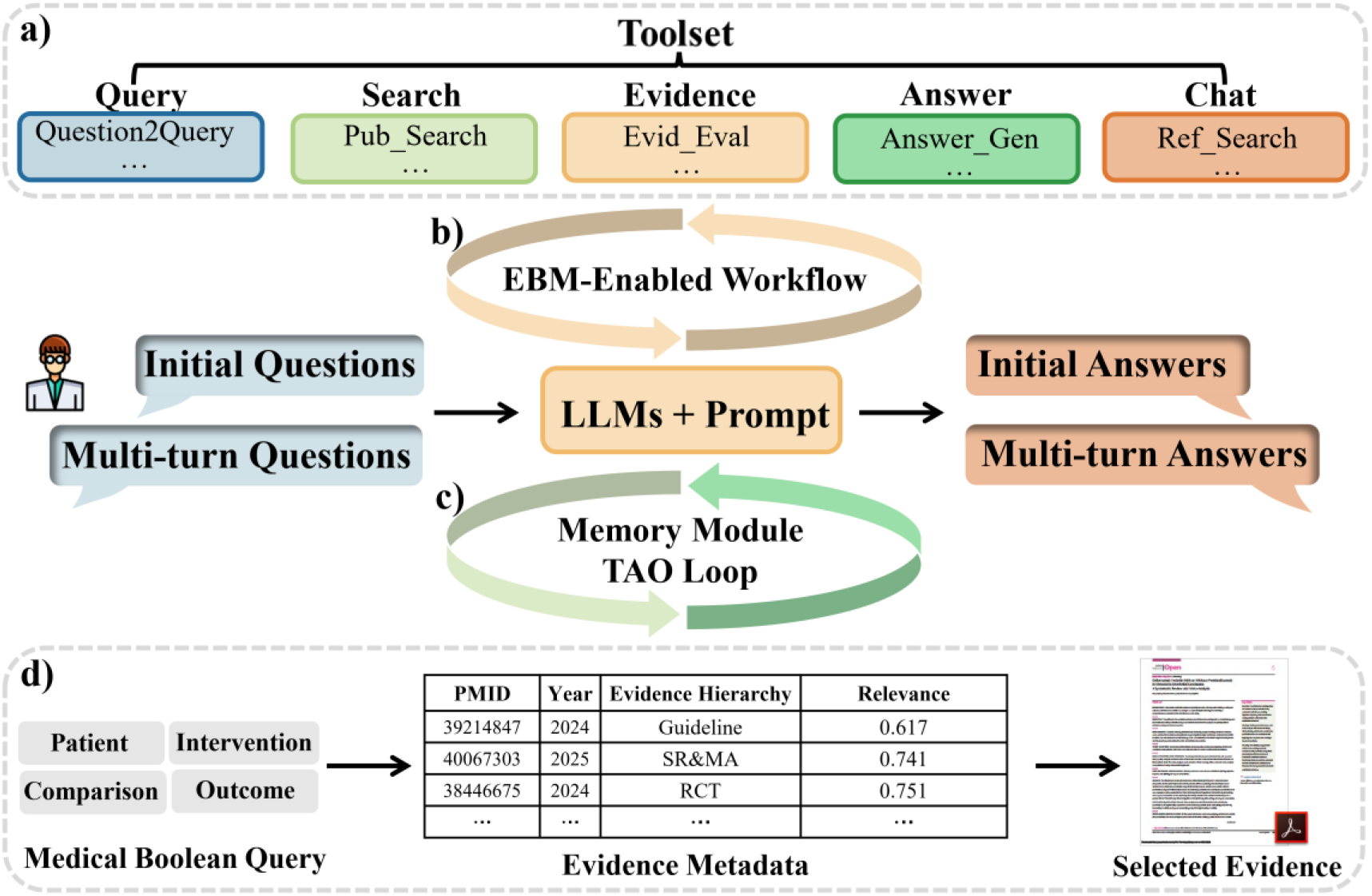
Conceptual and schematic representation of EBMChat. a) The toolset of EBMChat including five sections; b) EBM-enabled workflow for initial questions; c) The memory module and TAO loop for multi-turn questions. d) Contextual data generated in the iterative process, including queries, metadata containing information of studies and selected evidence.

The executor serves as the control center, managing both structured EBM-enabled workflow execution (**Figure 1b**) and dynamic Thought-Action-Observation (TAO) loops^27^ **(Figure 1c, eMethods**). The memory module is designed to store contextual information throughout multi-turn dialogues^28^.

Regarding the workflow of answering initial questions, firstly, a user submits a query. Secondly, EBMChat uses internal tools following structured workflow to explore the most appropriate evidence. In this iterative process, contextual information is also generated and stored in memory module, including queries, evidence metadata and selected evidence (**Figure 1d**). Thirdly, evidence-based answer is output by Answer_Gen tool based on the most suitable evidence. As for subsequent conversations, the previously stored contextual information in memory module will be called to ensure conversational coherence and consistency. Additionally, EBMChat autonomously reasons and selects tools to complete tasks through TAO loops. The example will be shown in the **Results**.

### Initial question preparation baseline and evaluation

We created 150 clinical questions by adapting the framework from Bitterman et al.^2^, comprising 75 general and 45 specific questions (Details in **eMethods)**. Three blinded raters (5,7 and 10 years of clinical experience in oncology) independently evaluated the accuracy for EBMChat and baselines. Each response was classified into one of three predefined categories— correct, inaccurate, or wrong, following the evaluation standard from Masanneck et al.^6^. Baselines include traditional academic (MEDRAG)^15^ and commercial (GPT-4.1+Web Search)^29^ methods.

### Evaluation of multi-turn dialogue

The evaluation of multi-turn dialogue performance comprised 45 tasks spanning three distinct types across 15 common cancer types. Three task types consist of presenting search result in specific category (Type I), answering questions by retrieving evidence from the bibliography of specific articles (Type II), and addressing new related questions based on the previous search results (Type III, all examples shown in **Figure 4** and **eFigure 1**). The baseline is GPT-4.1+Web Search. Task outcomes were dichotomized as successful or unsuccessful. Tasks were deemed successful when previous conversation is remembered and answer is correct, otherwise the task was considered unsuccessful (**eFigure 1**).

## Results

### Initial Q&A by EBM-enabled workflow

For an initial clinical question, EBMChat initiates EBM-enabled workflow that sequentially executes a series of tools: Question2Query, Query_Opt, PM_Search, Evid_Eval, and Answer_Gen (**Figure 2a**). Firstly, Question2Query converts the natural language question into an initial Boolean query. Inspired by the Wang’s work^30^, the conversion is realized by retaining the necessary keywords based on PICO framework^31^ and connecting by Boolean operators. Secondly, The initial query is then optimized by the Query_Opt tool, which expands term coverage by incorporating Medical Subject Headings (MeSH) terms 32 and adding keywords related to cancer staging where applicable^32^. Additionally, keywords about cancer staging were also expanded. The optimized query is constructed by combining the initial and modified parts with an “OR” operator (**Figure 2b**). Thirdly, search operation is performed based on the optimized query through PM_Search, returning a list of relevant literature with metadata including PMID, publication year, and title (**Figure 2c**). Meanwhile, The relevance of each item is ranked using cosine similarity^7^, and the evidence hierarchy is assigned based on publication type. Fourthly, Evid_Eval tool subsequently selects the most appropriate evidence by jointly considering publication year, evidence hierarchy, and relevance. In this instance, the systematic review and meta-analysis (SR/MA) with PMID 40067303 was selected. Fifthly, Answer_Gen tool is used to generate the final answer based on the evidence and RAG (**Figure 2d**). The example for a general question was illustrated in **eFigure 2**. following an identical EBM-enabled workflow.

**Figure 2.**
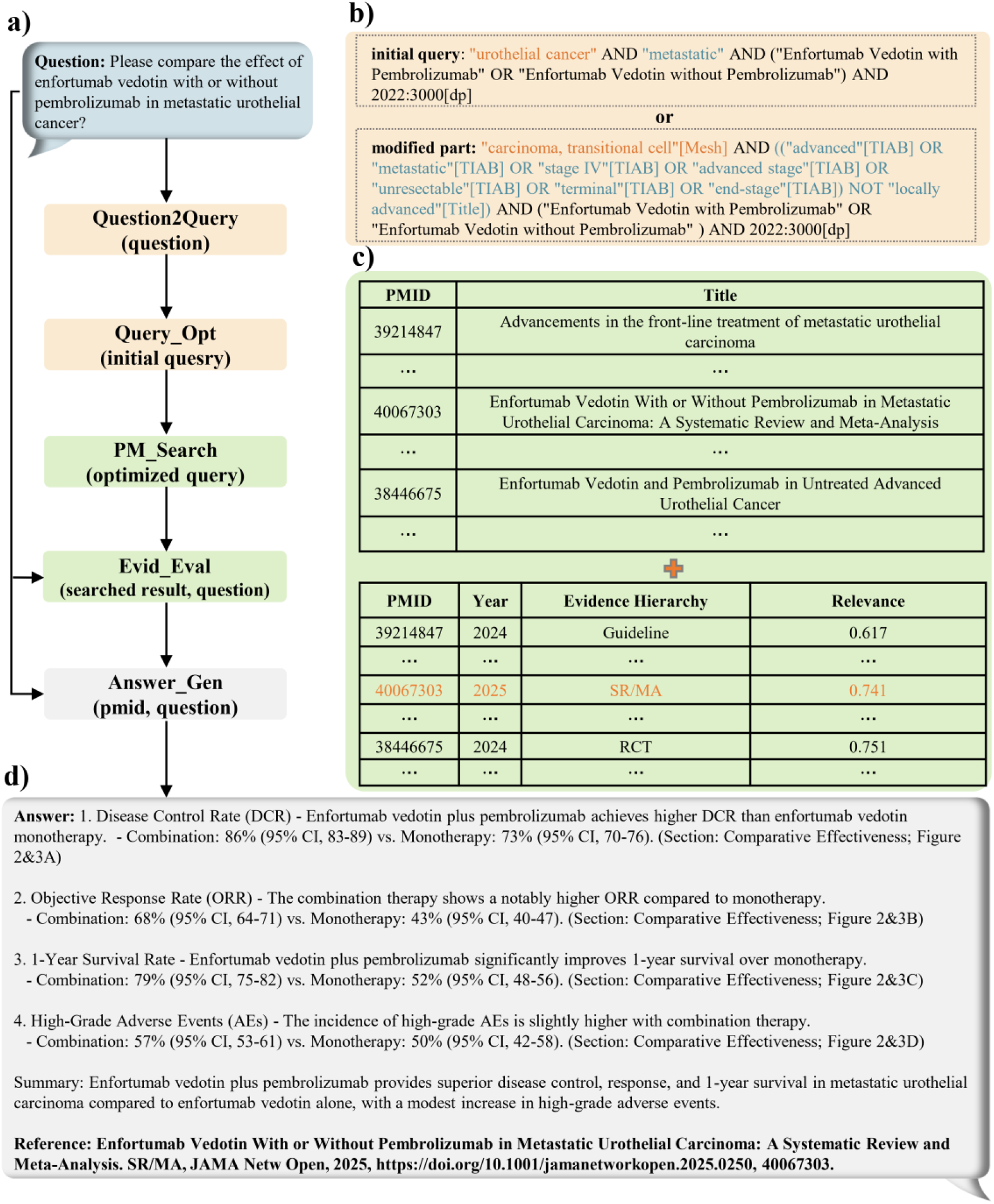
The example of initial Q&A by EBMChat. a) The specific question and agent’s evidence-based medicine workflow for answering the question; b) Question2Query tool converts the initial question into the initial query, then Query_Opt generate modified part of optimized query based on initial one (using colors to distinguish between different replacement parts). Two of them is connected by “OR” to form optimized query as the input for the next section; c) Pub_Search tool performs query search on PubMed and return the information of relevant studies, including PMIDs, years of publication, titles. Then relevance and evidence hierarchy are obtained by Evid_Eval when input is searched result and initial question. This tool then selects most suitable evidence for further answer generation process; d) Answer_Gen generates clinical answer (including citation) by RAG.

The mechanism of the Evid_Eval for assessing and retrieving the evidence is based on hierarchical algorithm (**Figure 3**). First, EBMChat selects the most appropriate evidence among studies of the same evidence hierarchy by incorporating timeliness and relevance (**Figure 3a**). In subsequent comparisons across publications with different evidence level, the system prioritizes higher-hierarchy evidence when relevance is comparable (**Figure 3b**). In the case above, although the RCT^33^ demonstrated the highest relevance, the SR/MA^34^ exhibited sufficiently high relevance while also possessing a higher evidence level than the RCT. Consequently, EBMChat ultimately selected the SR/MA as the evidence for subsequent answer-generation action. The advantage of evidence retrieval based on high-level evidence will be elaborated in **Discussion**.

**Figure 3.**
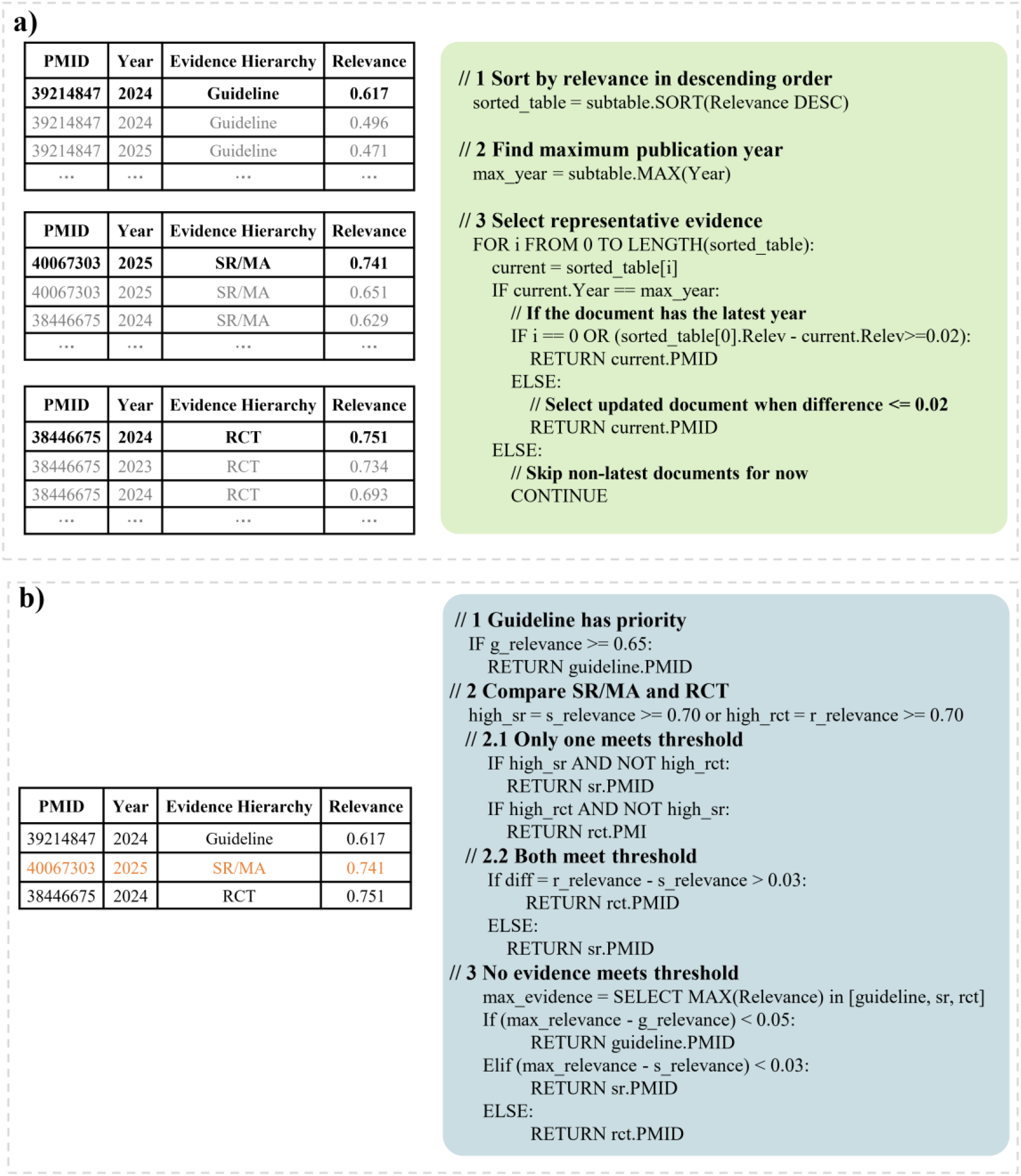
Mechanism and pseudo code of Evid_Eval for selecting most suitable evidence. a) With the same evidence hierarchy, the agent selected representative evidence (bold) based on both time and relevance for guidelines, SR/MAs and RCTs, respectively; b) Although the selected RCT has the highest relevance, selected SR/MA has higher evidence level and comparable relevance. Therefore, PMID 40067303 (Orange mark) was selected as the most appropriate evidence.

Besides evidence selection above, EBM-enabled query generation & optimization and evidence searching processes eliminate reliance on manually curated knowledge bases or knowledge graphs in previous method^35^. This end-to-end agent significantly lightens the workload for chatbot construction.

### Multi-turn dialogue through EBM-enabled workflow, memory module and TAO loop

**Figure 4** illustrates a multi-turn dialogue scenario where EBMChat addresses a continuous clinical question. The initial question (**Figure 4a**) prompts EBMChat to perform a standard EBM-enabled workflow mentioned in last section. However, based on the reference shown in the generated answer, it was obvious that the retrieved evidence—a guideline^36^ focused on radiotherapy for breast cancer—was not comprehensive enough to fully address the initial question. In response to this limitation, during the second-round dialogue (**Figure 4b**), the user explicitly requested to review the complete set of guideline information from the search result of from last dialogue. Through the TAO loop, EBMChat invoked the Table_Show tool to extract and display the stored search result from its memory module, which included detailed metadata such as PMIDs, publication years, titles, and relevance. In the third-round dialogue (**Figure 4c**), the user manually selected a specific article (PMID: 38101773)^37^ from the displayed list and requested an answer based on that evidence. Subsequently, EBMChat directly activated the Answer_Gen to generate a new response based on the user-specified evidence. During the fourth round (**Figure 4d**), driven by the user’s interest in adjuvant paclitaxel+trastuzumab, EBMChat automatically employed the Ref_Search tool to identify the most relevant reference as the new evidence within the bibliography from the guideline cited previously. Then it utilized the Answer_Gen to generate the deeper and more specific answer about the information of adjuvant therapy in detail.

**Figure 4.**
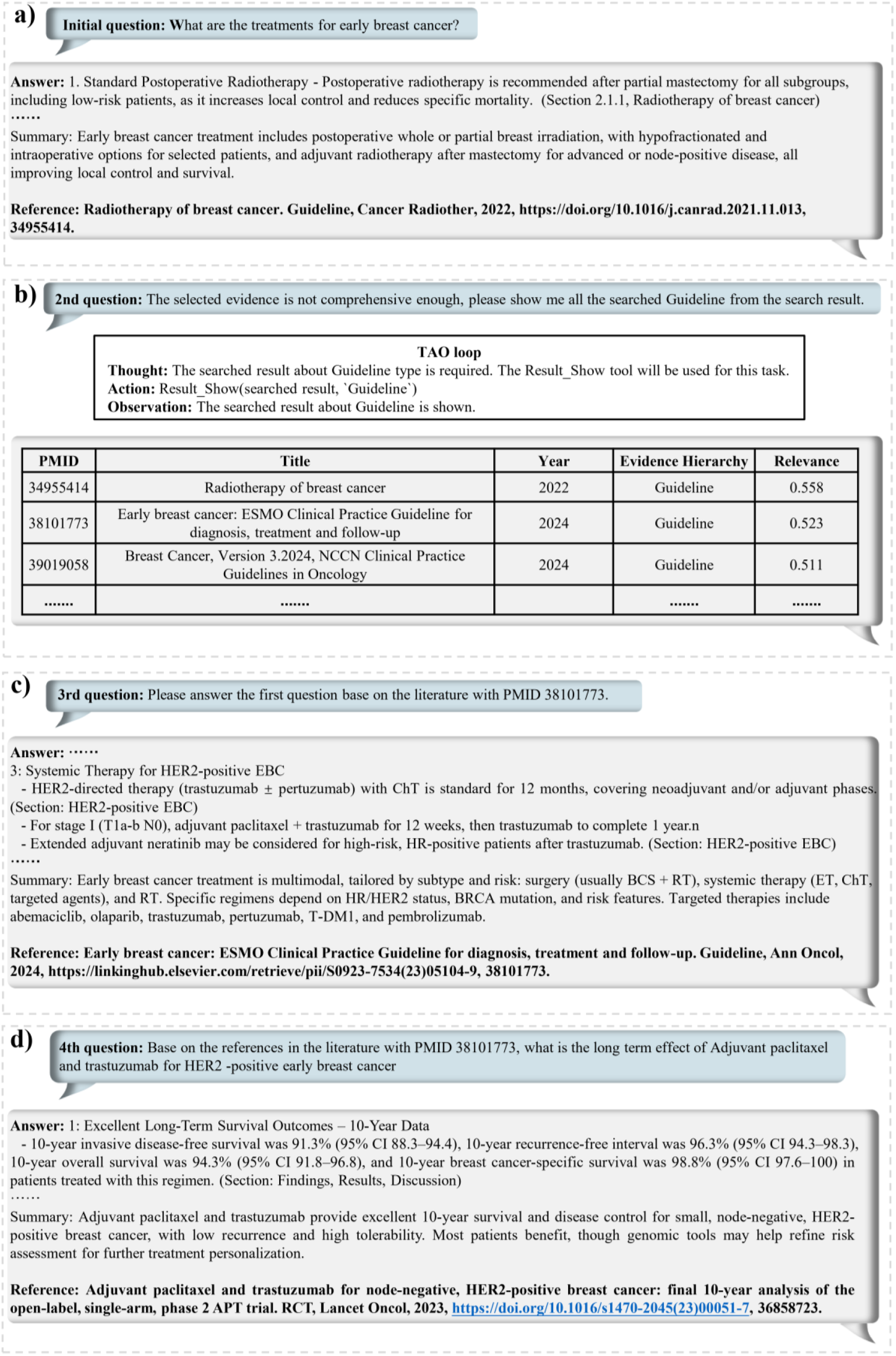
a) Initial Q&A about treatment for early breast cancer; b) The second round dialogue about the information of searched guidelines in the last round, Result_Show tool is used through thought-action-observation (TAO) loops; c) The third round about answering initial question based on user-specified evidence, Answer_Gen tool is used; d) The 4th round about answering new question based on the literature within the bibliography of the last selected evidence, Ref_Check and Answer_Gen tools are used through TAO loops.

Integrating with EBM-enabled workflow, EBMChat leverages its memory module to enhance performance in continuous dialogue tasks. The mechanism of this memory module is that the system can store not only the initial input or final output, but also the key intermediate information generated within entire workflows. More explicitly, EBMChat is able to retrieve contextual information according to users’ requirements during subsequent interactions, including the medical Boolean query, evidence metadata, and selected evidence. In addition, TAO loop ensures that EBMChat can autonomously reason and select tools to complete tasks (example in **Figure 4d**). Through the integration of EBM-enabled workflow, memory module and TAO loop, EBMChat demonstrates distinct advantages in user interactivity and problem-solving efficiency when handling multi-turn dialogues, which will be further explained in **Discussion**.

## Discussion

### Performance Comparison for initial questions

As shown in **Table 1**, EBMChat produced the highest proportion of accurate responses (89%) and the lowest incidence of wrong responses (1%), indicating its superior performance relative to other methods (all the Cohen’s kappa exceeded 0.91). The results for general and specific questions are detailed in **eTables 1 & 2**, respectively. Subsequently, we provide an in-depth analysis of the underlying reasons for the performance disparities among the different methods.

**Table 1.** Comparison result of 120 initial clinical questions between EBMChat and baselines. The classification includes Correct (ensuring comprehensive, timely and accurate), Inaccurate (with minor error) and Wrong (substantially incorrect, dangerous or misleading). MEDRAG-1 and MEDRAG-5 represent retrieve one or five studies as evidence for answer generation, respectively; Web Search represent use GPT-4.1 with its Web Search plugin provided by OpenAI.

| Method | Correct (%) | Inaccurate (%) | Wrong (%) | Cohen's kappa |
| --- | --- | --- | --- | --- |
| MEDRAG-1 | 40 | 36 | 24 | 0.91 |
| MEDRAG-5 | 78 | 18 | 4 | 0.92 |
| Web Search | 74 | 15 | 11 | 0.93 |
| EBMChat | 89 | 10 | 1 | 0.93 |

First, we compared EBMChat with MEDRAG-1. Although both methods base their responses on a single literature source, their evidence acquisition methodologies differ fundamentally. EBMChat adheres entirely to evidence-based medicine principles for evidence retrieval (**Figure 2**). While MEDRAG-1selects evidence through comparing the question’s relevance to titles and abstracts of the literature in PubMed Corpus^14^. Our analysis reveals that EBMChat holds significant advantages in three key aspects. Firstly, the proportion of high-evidence-hierarchy studies selected by EBMChat substantially surpassed MEDRAG-1 (**Figure 5a**). With similar relevance, RAG can leverage higher-hierarchy evidence to generate more accurate answers. For instance, regarding Question_**72** (a general query concerning prognosis for advanced **pancreatic** cancer), EBMChat retrieved a clinical guideline^38^ to offer the most comprehensive and authoritative solution, whereas the MEDRAG-1 approach identified an SR/MA^39^ whose answer disproportionately emphasized the situation about lung metastasis (**eFigure 3**). Secondly, EBMChat prioritizes evidence timeliness, while MEDRAG-1 frequently retrieves articles dating back a decade or more. As shown in **Figure 5b**, the evidence utilized in EBMChat consists of studies published within the past 5 years, whereas the references selected by the MEDRAG include a substantial number of sources that have become outdated. Outdated evidence is sometimes detrimental to clinical decision-making. Taking Question_9 as an example (**eFigure 4**), although both methods located literature of identical evidence levels, MEDRAG-1’s reliance on a 12-year-old source resulted in outdated treatment recommendations, potentially misleading clinical practice. Thirdly, EBMChat’s Query_Opt tool enabled EBMChat to retrieve a broader evidence base. Execution of the Query_Opt action significantly increased the median number of pertinent literature items retrieved per question from 267 to 693 (**Figure 5c**), thereby enhancing the likelihood of identifying optimal evidence (Question_81 as the example in **eFigure 5**). Hence, findings above demonstrated that when retrieving a single study as evidence for clinical questions, EBMChat consistently selected the most appropriate evidence to deliver superior answers.

**Figure 5.**
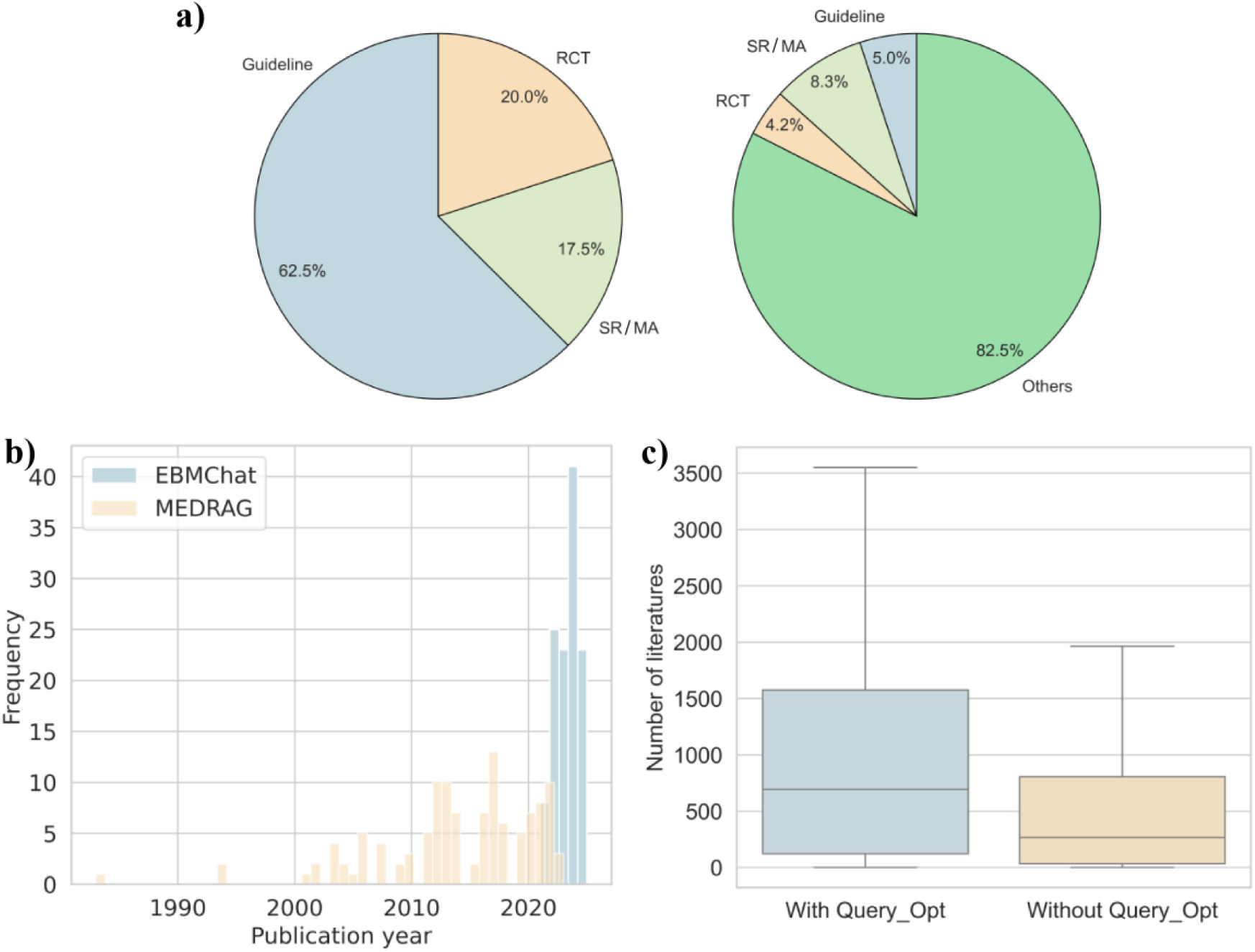
Three key advantages of EBMChat over MEDRAG. a) Pie charts showing the distribution of evidence levels for answers to 120 questions for EBMChat and MEDRAG, respectively; b) Histograms showing the publication year of 120 citations for EBMChat (blue) and MEDRAG (orange), respectively; c) Box plots about the distributions of the number of retrieved studies for 120 questions with (blue) and without (orange) Query_Opt tool.

Subsequently, we compared EBMChat against Web Search and MEDRAG-5. Unlike EBMChat and MEDRAG-1, the latter two methods synthesize answers using multiple studies as evidence sources. Despite of more retrieved sources, it is found that the optimal evidence may still not be included, which consequently resulted in lower accuracies for their answers. Moreover, our analysis revealed two further reasons for their overall worse performance. First, a single high-hierarchy evidence source from EBMChat is sufficiently systematic and comprehensive to encompass the findings of multiple papers, thereby synthesizing their findings to provide more reliable and quantifiable information. Question_115 exemplifies it (Q&As from 3 methods shown **eFigure** 6): EBMChat generated the answer based on single SR/MA^40^, whereas Web Search provided four cohort studies, and MEDRAG-5 offered one RCT and four cohort studies. It revealed that the answer from EBMChat covered more treatment details and quantified results than two other methods. This disparity arose because the SR/MA referenced by EBMChat incorporated all RCTs and cohort studies retrieved by the other two methods, plus five additional RCTs and over twenty relevant papers. Thus, EBMChat’s ability to leverage high-hierarchy evidence to yield more comprehensive and quantifiable answers even with fewer references. Second, after acquiring multiple studies, current RAG technology struggled to synthesize information efficiently and accurately, compared with EBM experts. Concretely, evidence retrieval from multi-source might result in omission of critical details, which was founded in previous work as well^6^. To clarify the problem, the case for Question_104 was shown in **eFigure 7**. Web Search method retrieved the same crucial RCT^41^ as EBMChat, as well as two other highly relevant studies. However, it omitted critical therapeutic information concerning the “sandwich sequence” (achieving highest 5-year survival rate). We posited this omission occurred because other retrieved studies lacked references to this specific regimen, leading LLMs to avoid mentioning this inconsistent across multiple sources. In contrast, the answer from EBMChat emphasized this treatment approach, due to the retrieval of optimal evidence.

Accordingly, compared with other methods, EBMChat’s superior performance across initial questions fundamentally stems from its EBM-enabled workflow to search and retrieve evidence.

### Performance Comparison for multi-turn dialogues

As for the evaluation of the multi-turn dialogue performance, EBMChat was able to successfully complete 93% of the tasks (successful for all Type I & III tasks, as well as 12 out of 15 Type II tasks). This achievement was attributable to EBMChat’s ability to recall critical information and use tool reasonably. In contrast, GPT-4.1+Web Search, the baseline, achieved a success rate of just 31% of multi-round dialogue tasks due to the absence of abilities mentioned above (**Table 2**). Subsequently, the performance differences across various task types will be analyzed separately.

**Table 2.**
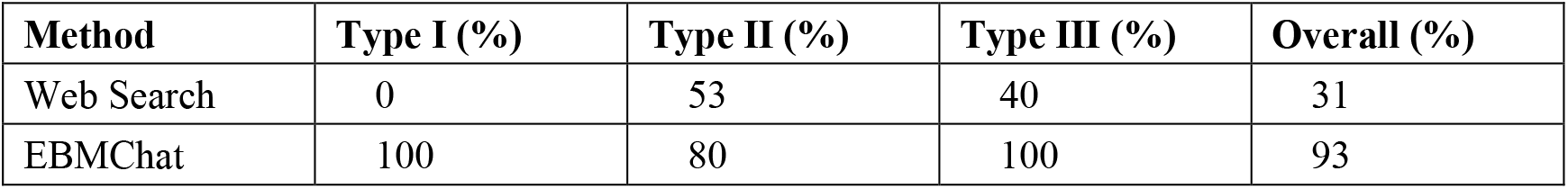
Success rates of 45 multi-turn clinical dialogue tasks between EBMChat and Web Search method.

| **Method** | **Type I (%)** | **Type II (%)** | **Type III (%)** | **Overall (%)** |
| --- | --- | --- | --- | --- |
| Web Search | 0 | 53 | 40 | 31 |
| EBMChat | 100 | 80 | 100 | 93 |

Regarding Type I tasks, EBMChat demonstrated a distinct advantage in user interactivity compared to the baseline. Actually, EBMChat empowers users to autonomously select the most appropriate evidence through the visualization of structured search results, thereby ensuring the possibility of correcting errors inherent in black-box systems. For instance, as shown in **eFigure 1a**, EBMChat returned 148 SR/MAs under user’s requirement, subsequently, users could select evidence from this broad space (case in **Figure 4**). Conversely, GPT-4.1+Web Search returned only 12. Therefore, this substantial difference reflected EBMChat’s user interactivity to recall conversation history and make active choices by themselves.

As for result of Type II & III tasks, the difference between EBMChat and baseline showed EBMChat’s enhanced problem-solving efficiency. In Type II tasks, EBMChat performs deeper exploration of fine-grained knowledge generated earlier in the conversation by recalling the bibliography information of selected evidence through the Ref_Search tool. This allows the system to efficiently access supporting references without initiating new searches. In contrast, the baseline failed due to LLMs’ hallucination^42^ (**eFigure 1b**). Meanwhile, in Type III tasks, EBMChat effectively addressed users’ new related questions based on originally retrieved search results, further elevating conversational efficiency. While GPT-4.1+Web Search could only redundantly start a new round of search and answer generation (**eFigure 1c**).

Accordingly, EBMChat—empowered by EBM-enabled workflow, memory module and TAO loops—established its user interactivity and problem-solving efficiency capability to in multi-turn dialogues, compared with commercial LLMs augmented with plugins.

## Conclusion

This study introduces EBMChat, an agent-based chatbot integrating evidence-based medicine principles and contextual conversation capabilities to answer clinical questions. The EBM-enabled workflow enables EBMChat to automatically generate and optimize medical queries, as well as classify and select optimal evidence. In this way, this end-to-end system can answer a wide range of clinical questions without prior laborious construction of vector databases or knowledge graphs. EBMChat achieved the highest accuracy (89%) on 150 clinical questions spanning 15 cancer types, outperforming traditional academic and commercial methods. This superior performance is attributable to its ability to identify optimal evidence by jointly considering timeliness, hierarchy, and relevance. Meanwhile, given EBM-enabled workflow, memory module and TAO loop, it obtains user interactivity and problem-solving efficiency to resolve more comprehensive or deeper contextual clinical problems. Consequently, EBMChat successfully completed 93% of multi-turn dialogue tasks, a rate substantially higher than that achieved by commercial LLMs with plugins (31%). Accordingly, our findings highlight that EBMChat can be a transformative AI agent to augment the performance of LLMs and the RAG in clinical decision-making. Moreover, EBMChat provides a robust framework for deeper integration of medicine principles and AI technologies into optimal patient care^43^, rather than direct clinical application of general-purpose AI tools.

## Data Availability

All data produced in the present study are available upon reasonable request to the authors

https://github.com/YiYuDL/EBMChat

## Author Contributions

Dr Yu had full access to all of the data in the study and takes responsibility for the integrity of the data and the accuracy of the data analysis.

Concept and design: Yi, Yaqin.

Acquisition, analysis, or interpretation of data: All authors.

Drafting of the manuscript: Yi, Yaqin.

Critical review of the manuscript for important intellectual content: Lingli, Yaqin.

Statistical analysis: Yi.

Obtained funding: Yi, Lingli.

Administrative, technical, or material support: Lingli, Yaqin.

