## Supplementary information for "Augmenting Large Language Models and Retrieval-Augmented Generation with an Evidence-Based Medicine-Enabled Agent System"

### Supplemental Online Content

#### EMethods

##### 1. Toolset

The toolset is divided into five sections: Database, Literature, Molecule, Search, and Dataframe. The tools in the same module handle similar molecular data processing tasks. The sections and representative are
shown below:

1.1 Query

**Question2Query:** This tool is designed to convert question in natural language into medical query. The conversion adheres to the principle of Patient (P), Intervention (I), Comparison (C), and Outcome (O), namely PICO.

**Query_Opt:** The tool is applied to transfer the input query into optimized query, ensuring that more relevant evidence can be found. In the transformation, keywords are preferentially mapped to Medical Subject Headings (MeSH) terms, and keywords related to tumor staging are also replaced using multiple synonymous terms.

1.2 Search

**PM_Search:** This tool performs a two-step operation. The first step is to search the query based on the input and return information about PMIDs, years of publication, titles, etc. The second step is to calculate the relevance and classify the evidence. Finally, all the information is saved as a table in the CSV format.

**Coc_Search:** The function of the tool is similar to that of the PM_Search, with the sole difference being the querying of the Cochrane Library database.

1.3 Literature

**Evid_Eval:** This tool implements evidence selection through a two-step process. In the first step, the most appropriate evidence is selected within the same evidence level, ensuring timeliness and relevance. In the second step, literatures across different evidence levels are compared, starting from guidelines and systematic reviews & meta-analyses (SR&MA). If the difference in relevance between a higher-level literature and the next lower-level literature is minimal, the higher-level literature is directly selected as the evidence. The output is the PMID ID of the best evidence.

1.4 Answer

Answer_Gen: The tool is designed to generate answers to original questions through a two-step process after receiving evidence PMIDs. In the first step, it searches local environment to determine whether the corresponding literatures associated with the provided PMIDs (possibly one literature or multiple) are available. If the required literature is not found locally, the tool autonomously identifies download links and retrieves the relevant papers (access by logging into the institutional intranet to facilitate seamless downloads is recommended). In the second step, the tool reads the PDF files of these papers and employs retrieval-augmented generation (RAG) techniques to synthesize responses tailored to the specific questions. The final output is the answer as well as citation.

1.5 Chat

Table_Gen: The principal capability of the tool is to present detailed information pertaining to

designated evidence types, leveraging the evidence search outcomes derived from prior interactions within the dialogue sequence.
Ref_Search: The function of the tool is to extract the most relevant references from the bibliography of a specified literature in response to a user's query. First, the reference section of the literature is extracted and compiled into a table. Subsequently, each article's title is assessed for relevance to the user's question through a scoring mechanism. Finally, the reference with the highest relevance score is selected and returned as the most pertinent source.

**2. Prompt**

Prompt engineering refers to the process of designing and refining input prompts to guide LLMs in generating desired outputs[1]. The primary function of prompt engineering is to enhance the model's performance by making the instructions clearer, more specific, and aligned with the task at hand.

The underlying principle of prompt engineering is based on how LLMs interpret language patterns and context. By manipulating the structure and content of the prompt, the model can be directed toward producing more relevant or specialized outputs[2]. This is achieved by providing the model with a well-defined context, precise formatting instructions, and clear task definitions.

In this work, the prompt is divided into several key components: PREFIX, FORMAT_INSTRUCTIONS, QUESTION_PROMPT, SUFFIX, FINAL_ANSWER_ACTION, and REPHRASE_TEMPLATE. Each of these components plays a specific role in shaping the interaction with the model:

The prompt of EBMChat is shown here:
FORMAT_INSTRUCTIONS = """You can only respond with a single complete "Thought, Action, Observation, ... Thought, Action, Observation ..." format
OR a single "Final Answer" format.
Complete format:
Thought: (reflect on your progress and decide what to do next)
Action: (the action name, should be one of [{tool_names}])
Observation: (the feedback or results obtained after executing an action)
OR
Thought: The task is completed.
Final Answer: (the final answer to the original input question)
"""

QUESTION_PROMPT = """
Answer the question below using the following tools:
{tool_strings}
Use the tools provided in the toolkit, and choose the most specific tool available for each action, please pay attention to the file format before making the choice.
Your final answer should contain all information necessary to answer all the questions and subquestions.

Attention: Importance from front to back, and plan your steps accordingly:

1. When confronted with an initial question, please sequentially executes the following tools: Question2Query, Query_Opt, PM_Search, Evid_Eval, and Answer_Gen.
2. In subsequent questions, when requested to show all the searched guidelines (or SR&MA or RCT), the system employs the "Table_Gen" tool to present detailed information regarding the specified evidence type.
3. In subsequent questions, when requested to show the most relevant question in the reference list of the specific literature, use "Ref_Search" tool to show the result.
4. In the subsequent questions, when requested to answer the question according to the most relevant question in the reference list of the specific literature, first use the "Ref_Search" tool to get the PMID, then use the "Answer_Gen" tool to answer the question.

.

Question: {input}

"""

SUFFIX = """Thought: {agent_scratchpad}"""

FINAL_ANSWER_ACTION = "Final Answer:"

REPHRASE_TEMPLATE = """Here, you are an assistant related to molecular data processing. You need to answer the provided question as best as you can, based on the provided solution guidance. The solution guidance follows the format "Thought, Action, Observation", where the 'Thought' statements reflect on your progress and decide what to do next. The rest of the text is information obtained to complement the reasoning sequence, and it is 100% accurate. Your task is to write an answer to the question based on the solution draft, and the following guidelines:

The text should have an educative and assistant-like tone, be accurate, follow the same reasoning sequence than the solution draft and explain how any conclusion is reached. When input includes "The task execution is completed, please check the error in this step.", please stop the reasoning above.

Question: {question}

Solution draft: {agent_ans}

Answer:

"""

**3. Thought-Action-Observation (TAO) loop**

Upon receiving the clinical question, EBMChat starts the Thought step to consider the task at hand. It then chooses the first tool from the toolset to promote the progression in the first Action. Once the tool is executed, the result and new information to refine the reasoning are returned as an Observation. After completing the loop of Thought, Action and Observation, the agent reassesses the Observation and determines the next loop. This could involve continuing the cycle by invoking other tools or concluding the process by delivering the final answer. These iterations enable EBMChat to methodically deconstruct intricate tasks into manageable steps.

**4. Generation strategy for 120 initial questions**

The 120 clinical questions are comprised of 75 general questions and 45 specific questions. Regarding general questions, the design of them is as follows: A represents treatments, diagnosis, or prognosis; B denotes the stage of cancer; and C indicates one of the 15 common cancer types (shown in the figure below). Through various combinations of A, B, and C, a total of 75 general questions were generated.

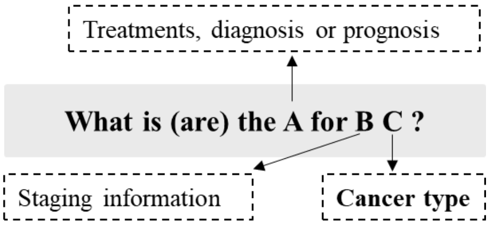

As for 45 specific questions, the design of the specific questions is as follows: A represents necessary conjunctions to make the question coherent; B denotes Targeted therapy, treatment comparisons or economic evaluations as the specific content; and C indicates one of the 15 common cancer types (shown in the figure below). The complete list of both general and specific questions can be found in Supplement 3.

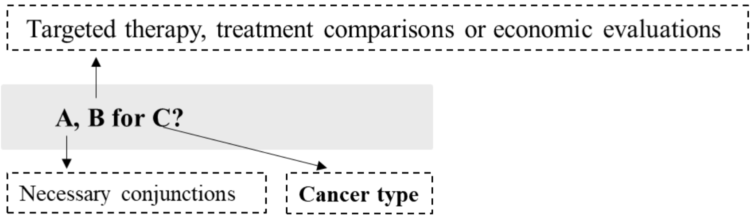

**a**

**Initial question:** What are the treatments for early breast cancer?

**Answer from EBMChat:** ……

**Answer from Web Search:** ……

**2nd question**: The selected evidence is not comprehensive enough, please show me all the searched SR/MA from the search result.

**Answer from EBMChat:**

| No. | Title | … |
|---|---|---|
| 1 | Adjuvant and neoadjuvant breast cancer treatments: A systematic review of their effects on mortality | … |
| 2 | Neoadjuvant and adjuvant treatment of patients with HER2-positive early breast cancer | … |
| 3 | Overall survival after mastectomy versus breast-conserving surgery with adjuvant radiotherapy for early-stage breast cancer: meta-analysis | … |
| … | … | … |
| 148 | Programmed death-ligand 1 (PD-L1) expression predicts response to neoadjuvant chemotherapy in triple-negative breast cancer: a systematic review and meta-analysis | … |

**Answer from Web Search:**

| No. | Title | … |
|---|---|---|
| 1 | Neoadjuvant immunotherapy and chemotherapy regimens for the treatment of high-risk, early-stage triple-negative breast cancer: a systematic review and network meta-analysis | … |
| 2 | Effectiveness of an Adjuvant Chemotherapy Regimen for Early-Stage Breast Cancer: A Systematic Review and Network Meta-analysis | … |
| 3 | The Omission of Anthracycline Chemotherapy in Women with Early HER2-Negative Breast Cancer—A Systematic Review and Meta-Analysis | … |
| … | … | … |
| 12 | Systematic review and meta-analysis of the efficacy of breast conservation therapy followed by radiotherapy in four breast cancer subtypes | … |

**b**

**3rd question:** Based on the bibliography of PMID 38101773, please tell me the information and direct link of the most relevant reference about "pregnancy-related information for breast cancer patients"

**Answer from EBMChat:**
PMID 34197218
Ref. 109: Pregnancy After Breast Cancer: A Systematic Review and Meta-Analysis. SR/MA, J Clin Oncol, 2021.
Direct link: https://doi.org/10.1200/JCO.21.00535

**Answer from Web Search:**
Reference 109: Lambertini M, Blondeaux E, Bruzzone M, et al. Pregnancy after breast cancer: a systematic review and meta-analysis. J Clin Oncol. 2021;39(29):3293-3305.
Direct link (via Journal of Clinical Oncology):
https://ascopubs.org/doi/10.1200/JCO.21.00662

**c**

**4th question:** Based on the initial search result, please tell me the information and direct link of the most relevant reference about "pregnancy-related information for breast cancer patients"

**EBMChat's TAO loop**
**Action_1:** Evid_Eval(searched result, initial question)
**Action_2:** Answer_Gen(pmid)

**Answer from EBMChat:**
1. Adjuvant Therapy Benefits - Reduces recurrence and improves survival in early-stage breast cancer by eradicating micrometastatic disease after surgery (Section: Adjuvant Systemic Therapy).
2. Adjuvant Therapy Risks - Associated with toxicities such as cardiotoxicity (anthracyclines), neuropathy (taxanes), and increased risk of secondary malignancies; may also impact quality of life (Section: Adjuvant Systemic Therapy—Toxicities).
3. Neoadjuvant Therapy Benefits - Downstages tumors, increases breast-conserving surgery rates, and provides early assessment of treatment response; may improve outcomes in locally advanced and some early-stage cancers (Section: Neoadjuvant Therapy).
4. Neoadjuvant Therapy Risks - Potential for disease progression if ineffective, and risk of overtreatment in some patients; similar toxicity profile to adjuvant therapy (Section: Neoadjuvant Therapy—Risks and Considerations).

Summary: Adjuvant and neoadjuvant breast cancer treatments improve survival and surgical options but carry risks of toxicity and potential overtreatment. Therapy choice depends on disease stage, tumor biology, and patient factors.

**Reference: Adjuvant and neoadjuvant breast cancer treatments: A systematic review of their effects on mortality. SRMA, Cancer Treat Rev, 2022, https://doi.org/10.1016/j.ctrv.2022.102375, 35367784.**

**Web Search process**
"adjuvant neoadjuvant breast cancer treatment benefits risks pubmed"

**Answer from Web Search:**
1. Adjuvant therapy after surgery reduces recurrence and improves survival but can cause side effects like low blood counts, heart issues, and other drug-specific risks.
2. Neoadjuvant therapy before surgery shrinks tumors and may allow breast conservation, but has similar side effects and a slightly higher risk of local recurrence.
3. Radiotherapy lowers local recurrence risk but may cause skin, heart, or lung problems.
4. Endocrine therapy benefits hormone receptor-positive patients but can increase bone, heart, or clotting risks.

Summary: Both adjuvant and neoadjuvant therapies are key for early breast cancer, improving outcomes but requiring careful selection based on tumor type and patient health.

**eFigure 1. Type I -III multi-turn dialogue tasks for EBMChat and Web Search methods.** a) type I task for presenting search result in specific category (SR/MA); b) Type II task for answering questions by retrieving evidence from the bibliography of specific articles (EBMChat gave the right reference link, while the link (in red color) from Web Search was wrong); c) Type III task for addressing new related questions based on the previous search results, EBMChat effectively addressed users' new related questions based on originally retrieved search results, further elevating conversational efficiency. While GPT-4.1+Web Search could only redundantly start a new round of search and answer generation.

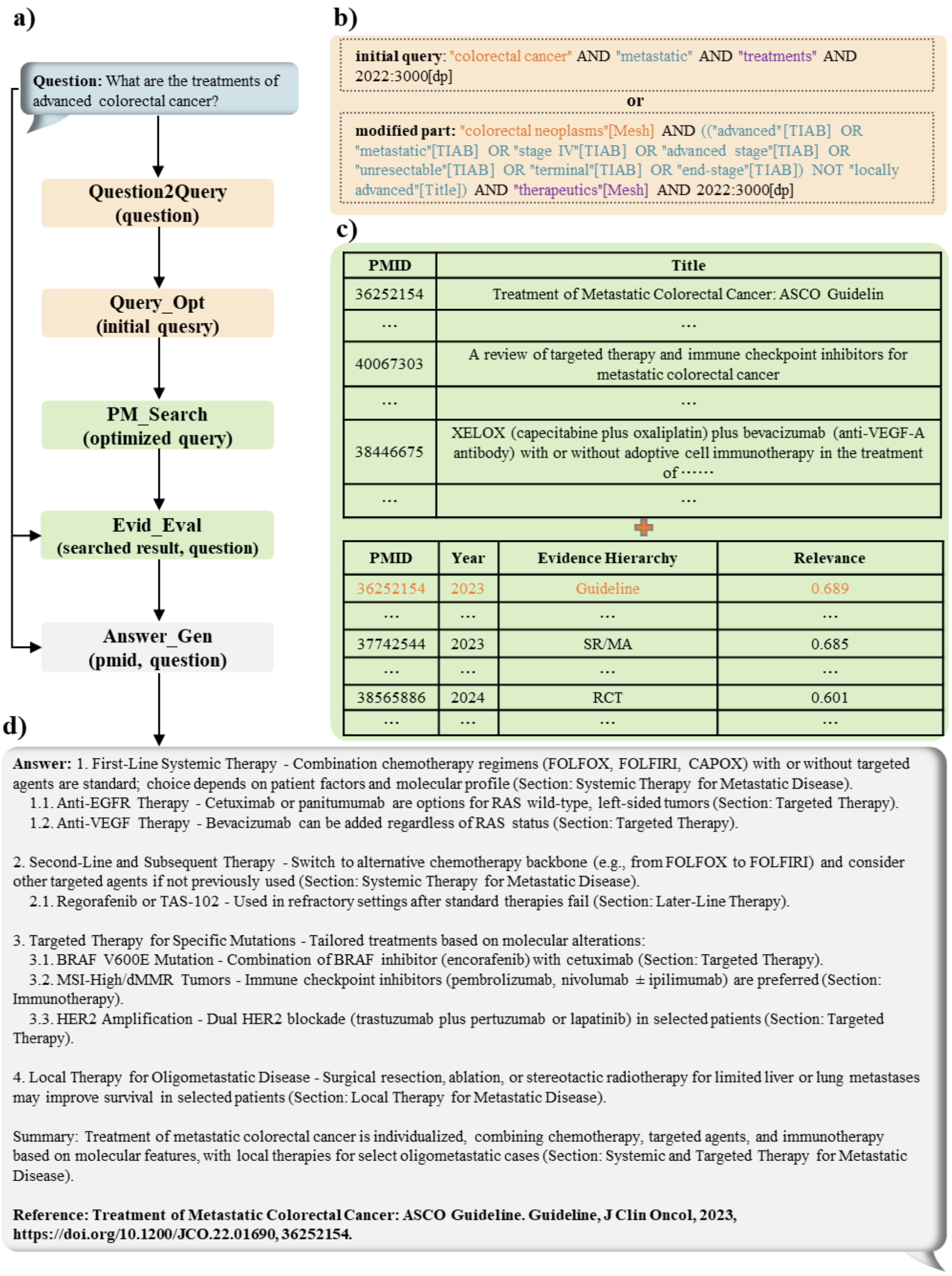

**eFigure 2. The example of initial Q&A by EBMChat.** a) The general question and agent's evidence-based medicine workflow for answering the question; b) Question2Query tool converts the initial question into the initial query, then Query_Opt generate modified part of optimized query based on initial one (using colors to distinguish between different replacement parts). Two of them is connected by "OR" to form optimized query as the input for the next section; c) Pub_Search tool performs query search on PubMed and return the information of relevant literatures, including PMIDs, years of publication, titles. Then relevance and evidence hierarchy are obtained by Evid_Eval when input is searched result and initial question. This tool then selects most suitable evidence for further answer generation process; d) Answer_Gen generates clinical answer (including citation) by RAG.

**Question:** What is the prognosis of metastatic pancreas cancer?

**Answer from EBMChat:**

1. Overall Survival (OS) - Median OS for metastatic pancreatic ductal adenocarcinoma (PDAC) remains poor, with recent first-line regimens achieving median OS of 9–11 months in clinical trials.
   - Sectioned in "MANAGEMENT OF ADVANCED DISEASE" and supported by NAPOLI-3 and other cited trials.

2. Prognostic Factors - Prognosis is influenced by ECOG performance status, nutritional status, bilirubin levels, and response to therapy.
   - Discussed in "MANAGEMENT OF ADVANCED DISEASE" and "Real-world prognostic factors for survival among treated patients with metastatic pancreatic ductal adenocarcinoma".

3. Impact of Treatment - FOLFIRINOX and NALIRIFOX regimens offer modest improvements in OS and progression-free survival (PFS) compared to gemcitabine-based therapies, but higher toxicity may limit use to fit patients.
   - Detailed in "First-line treatment" and NAPOLI-3 trial results.

4. Survival Rates - Despite advances, 5-year survival for metastatic PDAC remains extremely low (<5%), with most patients succumbing to disease within a year of diagnosis.
   - Implied throughout "MANAGEMENT OF ADVANCED DISEASE" and supported by referenced survival data.

Summary: Median OS for metastatic pancreatic cancer is 9–11 months with current therapies; prognosis is poor, influenced by performance status and treatment response, and 5-year survival remains below 5%.

**Reference: Pancreatic cancer: ESMO Clinical Practice Guideline for diagnosis, treatment and follow-up. Guideline, Ann Oncol, 2023, https://doi.org/10.1016/j.annonc.2023.08.009, 37678671.**

**Answer from MEDRAG:**

1. Prognosis by Metastatic Site - Patients with isolated pulmonary metastasis from pancreatic ductal adenocarcinoma (PDAC) have significantly better overall survival (OS) compared to those with hepatic, peritoneal, or locoregional metastases. (See: "Survival of patients with metastatic PDAC who have received surgical resection of the disease", "Oncologic outcomes of recurrent or metastatic PDAC patients according to the patterns of disease dissemination")

2. Survival Outcomes - Weighted median OS for patients with pulmonary recurrence after pancreatectomy is 34.7 months, versus 11.3 months for hepatic and 17.9 months for peritoneal metastases. For unresected PDAC with isolated lung metastasis, OS is 7.3 months. (See: "Survival of patients with metastatic PDAC who have received surgical resection of the disease")

3. Disease-Free and Survival After Recurrence - Median disease-free survival (DFS) and survival after recurrence (SAR) are also longer for pulmonary metastasis (DFS: 15.9 months, SAR: 16.5 months) compared to other sites. (See: "Oncologic outcomes of recurrent or metastatic PDAC patients according to the patterns of disease dissemination")

4. Prognostic Factors and Molecular Features - Pulmonary metastases are associated with more indolent disease and may have distinct molecular features (e.g., higher frequency of KRAS mutations, low ERCC1 expression), but the clinical impact of these findings is not fully understood. (See: "Molecular alterations in advanced PDAC according to the site of metastatic dissemination")

5. Limitations - Prognostic data are limited by heterogeneity in diagnostic methods, treatment regimens, and lack of randomized evidence. Not all pulmonary metastases had histological confirmation. (See: "Shortcomings and limitations of the present analysis")

Summary: Patients with isolated pulmonary metastasis from PDAC have significantly better prognosis than those with metastases to other sites, with longer OS, DFS, and SAR. Prognostic data are limited by study heterogeneity and incomplete molecular understanding.

**Reference: Metastases or primary recurrence to the lung is related to improved survival of pancreatic cancer as compared to other sites of dissemination. Results of a systematic review with meta-analysis. SR/MA, Eur J Surg Oncol, 2020, https://doi.org/10.1016/j.ejso.2020.06.013, 32753117.**

**eFigure 3. The difference between answer from EBMChat and MEDRAG-1 about evidence hierarchy.** EBMChat retrieved a clinical guideline to offer the most comprehensive and authoritative solution, whereas the MEDRAG-1 approach identified an SR/MA whose answer disproportionately emphasized the situation about lung metastasis

**Question:** What are the treatments of metastatic gastric cancer?

**Answer from EBMChat:**

1. First-Line Systemic Therapy (see "Principles of Systemic Therapy", "Targeted Therapies"):
Two-drug cytotoxic regimens (fluoropyrimidine + platinum) are preferred for most patients due to lower toxicity; three-drug regimens reserved for fit patients. Oxaliplatin is preferred over cisplatin for lower toxicity. Addition of targeted agents or immunotherapy is based on biomarker status:
   1.1. HER2-positive: Add trastuzumab to chemotherapy; add pembrolizumab if PD-L1 CPS $\geq 1$ (category 1 for CPS $\geq 1$) (see "Targeted Therapies: Trastuzumab, Pembrolizumab").
   1.2. HER2-negative, PD-L1 CPS $\geq 1$: Add immune checkpoint inhibitor (nivolumab, pembrolizumab, or tislelizumab) to chemotherapy (category 1 for CPS $\geq 5$) ("Targeted Therapies: Nivolumab, Pembrolizumab, Tislelizumab").
   1.3. CLDN18.2-positive: Add zolbetuximab to chemotherapy (category 1) ("Targeted Therapies: Zolbetuximab").
   1.4. MSI-H/dMMR: Pembrolizumab, dostarlimab, or nivolumab (± ipilimumab) are preferred options, independent of PD-L1 status ("Targeted Therapies: MSI-H/dMMR Tumors").
   1.5. NTRK fusion, BRAF V600E, RET fusion: Use corresponding targeted therapies (entrectinib/larotrectinib/repotrectinib, dabrafenib+trametinib, selpercatinib) if present ("Targeted Therapies").

2. Second-Line and Subsequent Therapy (see "Principles of Systemic Therapy", "Targeted Therapies"):
2.1. Preferred options: Ramucirumab + paclitaxel (category 1), single-agent docetaxel, paclitaxel, or irinotecan; FOLFIRI if not previously used; trifluridine/tipiracil for third-line or later.
2.2. HER2-positive: Fam-trastuzumab deruxtecan-nxki after progression on trastuzumab-based therapy.
2.3. Targetable mutations: Continue to use NTRK, BRAF, RET inhibitors as appropriate.
2.4. MSI-H/dMMR: Pembrolizumab, nivolumab+ipilimumab, or dostarlimab if not previously used.

3. Special Situations (see "Peritoneal Carcinoma as Only Disease"):
3.1. Peritoneal-only metastasis: Systemic therapy for $\geq 3$ months, then consider cytoreductive surgery + HIPEC for select patients with low peritoneal cancer index (PCI $\leq 10$) and no extraperitoneal disease; otherwise, continue systemic therapy or clinical trial.
3.2. Poor performance status (KPS <60% or ECOG $\geq 3$): Best supportive care only.

4. Palliative/Best Supportive Care (see "Unresectable Locally Advanced, Recurrent, or Metastatic Disease"):
4.1. Systemic therapy improves survival and quality of life compared to best supportive care alone.
4.2. Chemoradiation may be considered for unresectable, locally advanced disease if not previously received.

5. Treatment Selection Factors:
5.1. Therapy is tailored based on disease stage, biomarker status (HER2, PD-L1, CLDN18.2, MSI/MMR, NTRK, BRAF, RET), patient age, and performance status.
5.2. Participation in clinical trials is strongly encouraged.

Summary: Metastatic gastric cancer treatment is biomarker-driven, with first-line regimens based on HER2, PD-L1, CLDN18.2, and MSI/MMR status. Targeted agents and immunotherapy improve outcomes. Second-line options include ramucirumab-based regimens and further targeted therapies. Supportive care is essential for poor performance status.

**Reference: Gastric Cancer, Version 2.2025, NCCN Clinical Practice Guidelines In Oncology. Guideline, J Natl Compr Canc Netw, 2025, https://doi.org/10.6004/jnccn.2025.0022, 40341199.**

**Answer from MEDRAG:**

1. First-Line Chemotherapy - Combination regimens with platinum and fluoropyrimidine (e.g., FOLFOX, CAPOX, or cisplatin/5-FU) are standard first-line treatments for metastatic gastric cancer, improving survival and symptom control (Section: Systemic Therapy of Advanced Gastric Cancer).

2. Targeted Therapy for HER2-Positive Disease - Trastuzumab is added to chemotherapy for patients with HER2-positive tumors, showing improved outcomes compared to chemotherapy alone (Section: Targeted Therapy in Gastric Cancer).

3. Second-Line and Subsequent Therapy - Options include paclitaxel, irinotecan, or ramucirumab (alone or with paclitaxel), offering modest survival benefit after progression on first-line therapy (Section: Second-Line and Beyond Therapy).

4. Immunotherapy - Pembrolizumab or nivolumab may be considered for patients with PD-L1 positive, MSI-high, or dMMR tumors, especially after failure of standard therapies (Section: Immunotherapy in Gastric Cancer).

5. Palliative and Supportive Care - Symptom management, nutritional support, and palliative interventions are essential throughout the disease course to maintain quality of life (Section: Palliative Care in Gastric Cancer).

Summary: First-line treatment is platinum/fluoropyrimidine chemotherapy, with trastuzumab for HER2-positive cases. Second-line options include paclitaxel, irinotecan, or ramucirumab. Immunotherapy is used for select biomarker-positive patients. Palliative care is crucial for symptom control.

**Reference: Gastric cancer, version 2.2013: featured updates to the NCCN Guidelines. Guideline, J Natl Compr Canc Netw, 2013, https://doi.org/10.6004/jnccn.2013.0070, 23667204.**

**c**

**eFigure 4. The difference between answer from EBMChat and MEDRAG-1 about timeliness.**

Answer from MEDRAG represents a traditional, chemotherapy-centric approach, with targeted therapy limited to a single option (HER2) and no inclusion of immunotherapy; On the contrary, the other answer reflects a contemporary, biomarker-driven precision medicine model, featuring a rich array of targeted and immunotherapy options, with highly individualized treatment strategies.

**Question:** What is the marginal analysis result of radioembolization with Itrium-90 microspheres for liver cancers?

**Answer from EBMChat:**
1. Cost-Effectiveness in Intermediate-Advanced HCC – TARE with Yttrium-90 is generally cost-effective for intermediate-advanced hepatocellular carcinoma (HCC), showing favorable incremental cost-utility ratios (ICURs) compared to transarterial chemoembolization (TACE) and sorafenib. (Source: Abstract; Results; Discussion; Conclusion)

2. Marginal Analysis vs. TACE – TARE is associated with higher costs than TACE in most studies, but provides greater life-years gained (LYG) and quality-adjusted life-years (QALY) in intermediate-stage HCC. ICUR for TARE vs. TACE: ~$17,397/ QALY (US$ PPP). (Source: Results, "TARE versus TACE"; Table 3; Discussion)

3. Marginal Analysis vs. Sorafenib – TARE is often dominant (more effective, lower cost) or cost-effective compared to sorafenib in intermediate-advanced HCC, with ICURs ranging from dominant to ~$3,363/QALY (US$ PPP). (Source: Results, "TARE versus TKI"; Table 3; Discussion)

4. Budget Impact and Resource Use – TARE can reduce hospital resource use and overall costs, especially when compared to sorafenib, and may increase the number of patients eligible for curative treatments. (Source: Results, "Partial economic evaluations"; Discussion; Conclusion)

5. Influencing Factors – Cost-effectiveness varies by disease stage, comparator, country, and modeling assumptions. TARE is less cost-effective in early-stage HCC and when compared to ablative therapies. (Source: Results, Discussion)

Summary: Yttrium-90 TARE is cost-effective for intermediate-advanced HCC, especially versus sorafenib, with ICURs often below accepted thresholds. It offers greater life-years and QALYs than TACE, though at higher cost, and reduces hospital resource use compared to systemic therapy.

**Reference: Economic evaluations of radioembolization with Itrium-90 microspheres in hepatocellular carcinoma: a systematic review. SR/MA, BMC Gastroenterol, 2022, https://doi.org/10.1186/s12876-022-02396-6**

**Answer from MEDRAG:**
1. Cost-Utility Analysis -
a)Radioembolization showed a higher cost per quality-adjusted life year (QALY) compared to transarterial chemoembolization (TACE) but was considered cost-effective at willingness-to-pay thresholds ≥ $100,000/QALY (Section: Economic Evaluation).
b)Marginal cost-effectiveness ratios were influenced by tumor burden, Child-Pugh score, and baseline performance status (Section: Sensitivity Analysis).

2. Limitations of Analysis -
a)The study acknowledges uncertainties in long-term survival data for radioembolization, which may affect marginal cost estimates (Section: Discussion).
b)Real-world outcomes may vary due to differences in patient selection and treatment protocols (Section: Limitations).

Summary: The literature suggests radioembolization may be marginally cost-effective for liver cancers at higher willingness-to-pay thresholds, but uncertainties persist due to limited long-term data.

**Reference: Usefulness of Bremsstrahlung Images after Intra-arterial Y-90 Resin Microphere Radioembolization for Hepatic Tumors, Others, Nucl Med Mol Imaging, 2011, https://doi.org/10.1007/s13139-010-0066-4**

**eFigure 5. The difference between answer from EBMChat and MEDRAG-1 about the broadness of searched evidence.** The Query_Opt tool ensured EBMChat retrieved most appropriate evidence, whereas MEDRAG failed.

**a)** **Question:** What is the efficacy of streptozotocin in pancreatic neuroendocrine cancer?

**Answer from EBMChat:**

1. Indication and Patient Population - Streptozotocin (STZ)-based chemotherapy is primarily indicated for advanced, unresectable, or metastatic well-differentiated pancreatic neuroendocrine tumors (pan-NETs), especially in cases with rapid progression, high tumor burden, or symptomatic disease. (Section: Introduction; Characteristics of included studies and population)

2. Common Regimens - The most frequently used regimens are STZ combined with 5-fluorouracil (STZ-5FU) and STZ with anthracyclines (e.g., doxorubicin), administered as first-line or subsequent therapy. (Section: Characteristics of included studies and population)

3. Efficacy Outcomes (Advanced/Metastatic Disease) - Median progression-free survival (PFS) is 14.5 months (range: 3.9–31 months); median overall survival (OS) is 37.5 months (range: 10.9–69 months). Disease control rate (DCR) is 76.9%, and overall response rate (ORR) is 33%. (Section: Efficacy outcomes: Progression-free survival and overall survival; Objective response)

4. Efficacy in Perioperative Setting - In potentially resectable liver-only metastatic pan-NETs, median recurrence-free survival (RFS) is 25.1 months and median OS up to 108.2 months. DCR and ORR are higher in this setting (DCR 92.6%, ORR 63%). (Section: Efficacy outcomes: Progression-free survival and overall survival; Objective response)

5. Efficacy by Tumor Grade - Most benefit is observed in well-differentiated (G1-G2) pan-NETs; data on poorly differentiated or NEC are limited and less favorable. (Section: Characteristics of included studies and population)

Summary: STZ-based chemotherapy achieves a median PFS of 14.5 months and OS of 37.5 months in advanced pan-NETs, with DCR 76.9% and ORR 33%. Outcomes are best in well-differentiated tumors and perioperative settings.

**b)**

**Answer from MEDRAG-5:**

1. Treatment Regimen and Context
STZ was primarily administered to middle-aged patients (median age: 56) with advanced, well-differentiated pancreaticoduodenal NETs (96.4%), often with liver metastases. Intravenous regimens (weekly/bi-weekly) were common, particularly in Japan, and frequently combined with oral fluoropyrimidines like UFT or 5-FU, though 25% of patients received monotherapy.

2. Tumor Response
Per RECIST criteria, STZ yielded an objective response rate (ORR) of 28.2% and a disease control rate (DCR) of 86% in pancreaticoduodenal NETs. Responses included partial remission and prolonged stable disease, with some cases achieving long-term control.

3. Survival Outcomes
Median progression-free survival (PFS) was 11.8 months, and overall survival (OS) reached 38.7 months. Improved outcomes were associated with higher cumulative STZ doses (≥18.8 g), underscoring the role of dose intensity.

4. Safety Profile
Toxicity was generally mild; nausea (22.2%) and vomiting (13.0%) were most common. Grade 3 events were rare (11.1%), with no Grade 4 adverse effects or new-onset diabetes reported.

5. Comparative Efficacy
In a randomized trial, STZ plus capecitabine (CapStrep) achieved ORR 17% and DCR 86% in pancreatic NETs, with median OS of 26.7 months. Adding cisplatin (CapStrepCis) did not enhance efficacy (ORR 16%, OS 27.5 months) but increased toxicity.

Summary: STZ-based chemotherapy shows moderate efficacy (ORR ~28%, OS ~38.7 months) and manageable toxicity in advanced P-NETs. Fluoropyrimidine combinations are effective, but cisplatin adds no clinical benefit.

**c)**

**Answer from Web Search:**

1. STZ/5-FU: Standard Regimen

The STZ/5-FU combination remains a first-line standard for advanced pNETs, demonstrating an ORR of 25–38%, disease control rates of 70–80%, and median PFS of 12–23 months.

2. Treatment Line and Maintenance Impact

First-line use significantly prolongs median OS (up to 89 months vs. ~22 months later-line). Maintenance therapy with extended intervals enhances disease control and reduces toxicity.

3. Alternative Combinations with Doxorubicin

STZ plus doxorubicin shows marginally higher ORR (36–39%) in some studies, with biochemical regression >60% and median OS of 2–3 years.

4. Tumor Location and Efficacy

Tail-origin tumors respond better to STZ-based therapy (ORR 49%, PFS 31 months) compared to head-origin tumors (ORR 14%, PFS 16 months).

5. Safety and Contemporary Role

STZ carries risks of diabetes and nephrotoxicity but remains clinically valuable when newer agents are inaccessible or contraindicated.

Summary

STZ-based chemotherapy (with 5-FU ± doxorubicin) remains effective for advanced pNETs, particularly in first-line settings and tail-origin tumors. Maintenance strategies optimize outcomes, and STZ retains relevance despite newer alternatives.

**d)**

**Reference from EBMChat:**

The efficacy of streptozotocin in managing pancreatic neuroendocrine neoplasms - A systematic review. SR/MA, Cancer Treat Rev, 2025, https://doi.org/S0305-7372(25)00021-0, 39954330.

**Reference from MEDRAG-5:**

**Reference from Web Search:**

1. Streptozocin chemotherapy for advanced/metastatic well-differentiated neuroendocrine tumors: an analysis of a multi-center survey in Japan. Cohort, J Gastroenterol, 2015, https://doi.org/10.1007/s00535-014-1006-3, 25348496.
2. Streptozocin-Based Chemotherapy in Patients with Advanced Neuroendocrine Neoplasms--Predictive and Prognostic Markers for Treatment Stratification. Cohort, PLoS One, 2015, https://doi.org/10.1371/journal.pone.0143822, 26630134.
3. Streptozocin and 5-Fluorouracil for the Treatment of Pancreatic Neuroendocrine Tumors: Efficacy, Prognostic Factors and Toxicity. Cohort, Neuroendocrinology, 2016, https://doi.org/10.1159/000439086, 26279284.
4. Streptozocin/5-fluorouracil chemotherapy is associated with durable response in patients with advanced pancreatic neuroendocrine tumours. Cohort, Eur J Cancer, 2015, https://doi.org/S0959-8049(15)00309-3, 25935542.

**eFigure 6. Answers from EBMChat (a), MEDRAG-5 (b), Web Search (c) and references (d), respectively.** The answer from EBMChat is more comprehensive and includes valuable information from more studies, than answer from two other methods (The comprehensiveness of the answer is marked in red).

**a)** **Question:** what is the sequencing of checkpoint or BRAF/MEK inhibitors on brain metastases in melanoma?

**Answer from EBMChat:**

1. Treatment Sequencing Impact – The sequence of immune checkpoint inhibitors (ICIs) and BRAF/MEK inhibitors significantly affects brain metastases–free survival (BMFS) in unresectable metastatic BRAF V600-mutant melanoma.

— Patients receiving ICIs (ipilimumab + nivolumab) first, followed by BRAF/MEK inhibitors (encorafenib + binimetinib), had longer BMFS compared to the reverse sequence.

2. SECOMBIT Trial Results – At 60 months, BMFS rates were:

— 56% for first-line BRAF/MEK inhibitors (Arm A),

— 80% for first-line ICIs (Arm B),

— 85% for the "sandwich" regimen (Arm C: short BRAF/MEK induction, then ICIs, then BRAF/MEK at progression).

3. Characteristics of Brain Metastases – Brain metastases after first-line BRAF/MEK inhibitors were more frequent and had poorer features (e.g., more leptomeningeal disease, symptomatic lesions, higher need for whole brain radiotherapy and steroids).

4. Overall Survival (OS) – OS was similar across arms, regardless of the number of metastatic sites, but survival after brain metastases diagnosis was longer in patients who received ICIs first.

5. Clinical Implication – Initiating treatment with ICIs, or using a "sandwich" approach, may better prevent or delay brain metastases than starting with BRAF/MEK inhibitors. (Section: "Abstract", "Discussion")

Summary: Starting with immune checkpoint inhibitors, or using a "sandwich" regimen, provides superior brain metastases–free survival in BRAF V600-mutant metastatic melanoma compared to first-line BRAF/MEK inhibitors, with additional benefits in brain metastases characteristics and post-metastasis survival.

**b)**

**Answer from MEDRAG-5:**

1. First-line PD-1/CTLA-4 Inhibitors Superior: First-line ipilimumab-nivolumab provides high intracranial response rates (75%) and durable progression-free survival (median 41.6 months) for melanoma brain metastases (MBM), whereas first-line BRAF/MEK inhibitors show a much shorter PFS (~5.6 months) and frequent central nervous system (CNS) progression despite similar initial response rates.

2. Poor Efficacy of Second-line Immunotherapy: Using ipilimumab-nivolumab after progression on BRAF/MEK inhibitors results in very low intracranial response rates (4.8%) and a short median PFS (1.3 months), indicating significant cross-resistance.

3. Clinical Recommendation: Sequence Matters: For BRAFV600-mutant MBM, first-line combination immunotherapy (ipilimumab-nivolumab) is preferred due to more durable responses; BRAF/MEK inhibitors are reserved for cases requiring rapid symptom control or when immunotherapy is contraindicated.

4. Prognostic Factors and Real-World Data: Prognosis with BRAF/MEK inhibitors is worse in patients with factors like high LDH, poor ECOG performance status, and multiple metastatic sites. Real-world data confirms shorter PFS and a higher risk of brain progression with targeted therapy compared to immunotherapy.

Summary: First-line combination immunotherapy (PD-1/CTLA-4 inhibitors) offers superior and durable intracranial control for BRAFV600-mutant melanoma brain metastases compared to BRAF/MEK inhibitors. Using immunotherapy after the failure of targeted therapy is largely ineffective due to cross-resistance, underpinned by the IPRES signature. Upfront immunotherapy is the recommended strategy unless rapid symptom relief is urgently needed.

**c)**

**Answer from Web Search:**

1. The SECOMBIT trial demonstrated that initiating treatment with immune checkpoint inhibitors (ICIs) like ipilimumab and nivolumab, followed by BRAF/MEK inhibitors upon progression, resulted in significantly longer freedom from new brain metastases (80% at 60 months) compared to the reverse sequence (56%).

2. A multicenter cohort study supports that adding anti-PD-1 therapy to BRAF inhibitor-based treatment improves real-world survival and delays the onset of brain metastases.

3. Conversely, research indicates that brain metastases progressing on BRAF-MEK inhibitors may develop resistance to subsequent ICI therapy (ipilimumab-nivolumab), which is associated with the Innate PD-1 Resistance Signature (IPRES).

Summary: The evidence suggests that a treatment sequence starting with immune checkpoint inhibitors before switching to BRAF/MEK inhibitors is more effective for preventing or delaying brain metastases in patients with BRAFV600-mutant melanoma and may improve overall survival. However, treatment decisions should still be individualized based on patient factors and disease characteristics.

**d)**

**Reference from EBMChat:**

Sequencing of Checkpoint or BRAF/MEK Inhibitors on Brain Metastases in Melanoma. RCT, NEJM Evid, 2024, https://doi.org/10.1056/EVIDoa2400087, 39315864.

**Reference from MEDRAG-5:**

**Reference from Web Search:**

1. Sequencing of Checkpoint or BRAF/MEK Inhibitors on Brain Metastases in Melanoma. RCT, NEJM Evid, 2024, https://doi.org/10.1056/EVIDoa2400087, 39315864.
2. Melanoma brain metastases that progress on BRAF-MEK inhibitors demonstrate resistance to ipilimumab-nivolumab that is associated with the Innate PD-1 Resistance Signature (IPRES). Cohort, J Immunother Cancer, 2021, https://doi.org/10.1136/jitc-2021-002995, 34625515.
4. Addition of anti-PD-1 immunotherapy to BRAF inhibitor-based targeted therapy improves real-world survival and delays brain metastases in patients with BRAF(V600)-mutant advanced melanoma: a multicenter cohort study. Cohort, MedComm (2020), 2025, https://doi.org/10.1002/mco2.70102, 39968494.

**eFigure 7. Answers from EBMChat (a), MEDRAG-5 (b), Web Search (c) and references (d).** The answer from Web Search method retrieved the same RCT evidence as the one from EBMChat. However, it omitted critical therapeutic information, since two other literatures don't include this information.

**eTable 1 Comparison result of 75 general clinical questions between EBMChat and baselines.**

| Method | Correct (%) | Inaccurate (%) | Wrong (%) |
|---|---|---|---|
| MEDRAG-1 | 41 | 33 | 25 |
| MEDRAG-5 | 82 | 17 | 1 |
| Web Search | 80 | 11 | 9 |
| EBMChat | 85 | 15 | 0 |

**eTable 2 Comparison result of 45 specific clinical questions between EBMChat and baselines.**

| **Method** | **Correct (%)** | **Inaccurate (%)** | **Wrong (%)** |
|---|---|---|---|
| MEDRAG-1 | 38 | 40 | 22 |
| MEDRAG-5 | 71 | 19 | 10 |
| Web Search | 64 | 21 | 14 |
| EBMChat | 96 | 2 | 2 |
